# Local measures enable COVID-19 containment with fewer restrictions due to cooperative effects

**DOI:** 10.1101/2020.07.24.20161364

**Authors:** Philip Bittihn, Lukas Hupe, Jonas Isensee, Ramin Golestanian

## Abstract

Many countries worldwide that were successful in containing the first wave of the COVID-19 epidemic are faced with the seemingly impossible choice between the resurgence of infections and endangering the economic and mental well-being of their citizens. While blanket measures are slowly being lifted and infection numbers are monitored, a systematic strategy for balancing contact restrictions and the freedom necessary for a functioning society long-term in the absence of a vaccine is currently lacking. Here, we propose a regional strategy with locally triggered containment measures that can largely circumvent this trade-off and substantially lower the magnitude of restrictions the average individual will have to endure in the near future. For the simulation of future disease dynamics and its control, we use current data on the spread of COVID-19 in Germany, Italy, England, New York State and Florida, taking into account the regional structure of each country and their past lockdown efficiency. Overall, our analysis shows that tight regional control in the short term can lead to long-term net benefits due to small-number effects which are amplified by the regional subdivision and crucially depend on the rate of cross-regional contacts. We outline the mechanisms and parameters responsible for these benefits and suggest possible was to gain access to them, simultaneously achieving more freedom for the population and successfully containing the epidemic. Our open-source simulation code is freely available and can be readily adapted to other countries. We hope that our analysis will help create sustainable, theory-driven long-term strategies for the management of the COVID-19 epidemic until therapy or immunization options are available.

## Main

While the levels of daily new infections with COVID-19 around the world are still at an all-time high [1], many countries that were hit early on by the epidemic have demonstrated that its control is possible through population-wide contact bans and radical restrictions on public and economic life [1, 2, 3, 4, 5, 6], which were deemed necessary despite their socioeconomic cost. However, after the decline of infection numbers, these countries now face an equally important task, which is restoring socioeconomic life to sustainable levels without risking the resurgence of the epidemic.

Although some aspects of public life in these countries have already resumed, it has so far been possible to maintain relatively low, stable infection numbers, suggesting that the basic reproduction number is in the vicinity of 1. Given the volatility of a situation with *R* ≈ 1, governments must now prepare to systematically manage the epidemic with non-pharmaceutical interventions until medication or a vaccine is available. If the further reopening leads to a basic reproduction number > 1 even under the continued enforcement of hygiene rules, effective contact tracing and quarantine [7, 8], it is essential that such control strategies are capable of containing new outbreaks with minimal restrictions, in order not to stall the recovery process.

A natural idea for such more fine-grained approaches is to give more control to local authorities and employ regional containment measures only when necessary. In Germany, for example, many counties have not seen any new infections with the last 7 days [9], rendering control measures unnecessary until infections are reintroduced from outside (given sufficient testing).

### An adaptive local containment strategy

We set out to investigate a family of containment strategies for COVID-19 which aim at giving communities without substantial infection levels as much freedom as possible at any given time while trying to contain local outbreaks. We then determine the ability ob these strategies to avoid harsh contact restrictions for the average person in the population. Our proposed rules are based on critical infection thresholds that trigger restrictions in individual counties. To this end, we obtained the county structure of five countries/states, namely Germany, England, Italy, New York State and Florida, to set up a mathematical model composed of individual sub-populations and numerically simulate the future evolution of the epidemic starting from current case numbers. While the free spread of diseases through such subdivided populations [10, 11, 12] or networks [13, 14] has been investigated theoretically, we are not aware of previous studies that have considered the regional structure of specific countries and its relation to local containment measures in the context of COVID-19.

We use an extended SIR model that differentiates between internal contacts within a region and cross-regional contacts with the general population (Fig. 1a). Because of low infection numbers in each region, it is necessary to use a stochastic model with discrete infection events [15] (see Appendix for exact model definition). We assume that the reproduction number *R*_0_ in the overall population in the absence of stringent restrictions is slightly above 1. This is a reasonable assumption given that many countries have been successful in maintaining low infection numbers even after lifting their most strict lockdown measures [6], and while some restrictions remain to be removed, general hygiene and distancing measures are unlikely to be given up in the near future. However, to cover a wider range of parameters, we also simulated higher values of *R*_0_ up to *R*_0_ ≈ 1.7. The majority of an individual’s contacts take place in the local sub-population (small arrows in Fig. 1a), while some proportion of contacts with the general population (large arrows in Fig. 1a; parameterized by the *leakiness ξ*) can potentially lead to the spreading of the disease across sub-population boundaries.

**Figure 1.**
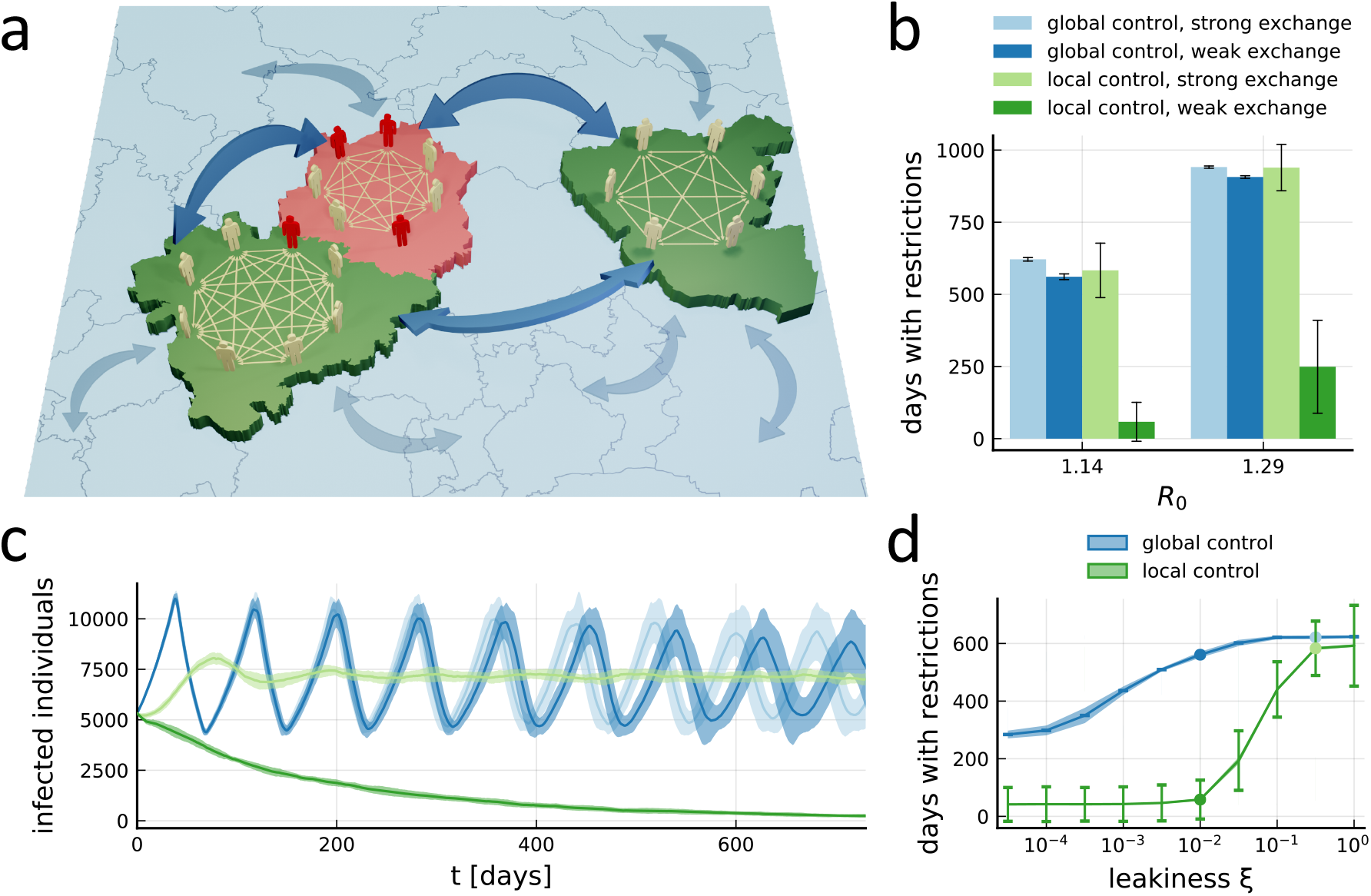
Freedom from restrictions through local containment measures. **a**, Illustration of core model ingredients: Individuals have contacts with random individuals inside their local sub-population (small arrows), which is defined, e.g., by a county. A certain proportion of contacts takes place across sub-population boundaries with random individuals from the whole population (large arrows). This *leakiness ξ* is defined such that the total number of contacts an individual has per unit time is a *ξ*-independent constant, which determines the reproductive number *R*_0_. If the number of infected individuals (red) in a sub-population exceeds a certain threshold, the sub-population enters a temporary local lockdown that lowers *R*_0_ and infection numbers can decline (red county). A precise model definition is available in the Appendix. **b**, Number of days the average person in the population will have to spend in lockdowns within the next 5 years, using the county structure and current active case numbers from Germany, with counties further split up into sub-populations of a maximum population of 200,000 (see Figs. 2, 3 for details). Numbers are shown for high (*ξ* = 32%) or low (*ξ* = 1%) leakiness between sub-populations, and for the local containment strategy outlined in panel A or an analogous population-wide (‘global’) strategy, *τ*_safety_ = 21 days, *θ* = 10 : 100,000. The four cases are shown for two different values of *R*_0_. Error bars indicate total standard deviation across members of all populations in 20 realizations. **c**, Time evolution in the first two years of the total number of infected individuals in the entire population for the four cases shown in panel B for the lower value of *R*_0_. Shading indicates standard deviation across 20 realizations. **d**, Restriction time for the full spectrum of leakiness values. The four cases shown in panels B and C (lower value of *R*_0_) are marked by dots of the same color. See Supp. Figs. 2, 4, 5, 6, 7, 8, 9 for additional parameters. Error bars indicate standard deviation across members of the population in a single simulation (averaged across 20 realizations), shading around lines indicates standard deviation of the average across realizations.

To give the population as much freedom as possible, local sub-populations only activate local restrictions (red county in Fig. 1a) whenever infection numbers cross a certain threshold *θ*, specified as number of infections per 100,000 inhabitants. For our main investigation, we assumed that the effect of local restrictions on the contact rate is similar to that of the recent population-wide lockdown measures, which allowed us to extract the corresponding reproduction number during the lockdown, *R*_*l*_, for each country directly from the available infection number data (see Appendix and Supp. Fig. 1). Similar values in the range between 0.77 and 0.79 were found for Germany, England, Italy, and New York State, which have had low infection numbers since the measures were imposed, and *R*_*l*_ ≈ 0.9 was found for Florida, which has had a resurgence after the initial period of control.

Since authorities have to rely on reported infection numbers, we make the conservative assumption that local restrictions can only be activated with a delay of *τ* = 14 days, which is made up of at least three contributions: First, case numbers are themselves reported with considerable delay because most testing only happens after the onset of symptoms and the administrative reporting process needs time. Second, in the presence of a constant background noise of fluctuating infection levels, heightened case numbers can only be detected with statistical significance after a certain amount of time [5]. Finally, a local lockdown needs to be ordered by the responsible government agencies, which adds additional reaction time. On the other hand, when local restrictions finally bring infection numbers down, they are only lifted after a period *τ*_safety_, which can be extended at will to allow for stringent control of the epidemic in a region.

### Local restrictions can be more effective than population-wide measures

With the rules set up as specified above, we could then compare the impact of such local containment strategies to equivalent population-wide strategies with the same parameters, where a population-wide lockdown reducing the reproduction number to *R*_*l*_ is activated once case numbers in the whole population reach *θ* (with identical delays *τ* and *τ*_safety_). To characterize this impact, we simulated the future evolution of the epidemic in the next 5 years and recorded the average time individuals in the population experience restrictions imposed during supra-threshold infection levels.

Assuming a high leakiness that leads to a strong exchange of infections between regions, we find that population-wide strategy (‘global control’) and local measures (‘local control’) perform similarly on average (Fig. 1b). On the one hand, this confirms that individuals on average are not worse off under thoroughly enforced local strategy than they would have been under a centrally managed lockdown strategy with equivalent parameters. However, it also means that the avoidance of unnecessary restrictions on the local level during times of low infection levels is a deception: On average, they will have to suffer the same amount of restrictions in both cases. This can also be seen in the evolution of overall infection numbers (Fig. 1c): While the fingerprint of alternating phases of population-wide freedom and population-wide lockdown can be clearly seen for the population-wide strategy, the local strategy with the same strong exchange between regions settles at very similar infection levels. Therefore, it does not come at a surprise that a similar amount of restrictions is required to manage them. Note that the local strategy at high leakiness, while not changing the average restriction time, introduces a large variability across members of the population (Fig. 1b), so some individuals will actually see an increase in restrictions compared to the population-wide strategy.

For a lower leakiness, stark differences appear: The lower exchange between sub-populations has almost no impact on the population-wide strategy, neither in terms of the average lockdown time required (Fig. 1b) nor in terms of disease dynamics (Fig. 1c). In contrast, the local strategy can control the epidemic while imposing restrictions for a substantially lower amount of time (Fig. 1b). The evolution of infection numbers in this case shows a steady decline towards complete extinction of the disease (Fig. 1c). This is the result of a cooperative effect between the local measures in different regions: Because of the targeted way in which local measures are applied, they have the chance of rendering individual regions disease-free by the end of the local lockdown or shortly afterwards through extinction [12], initiating periods of quiescence without the need for restrictions. This happens at a faster rate than infections can be reintroduced through cross infections for sufficiently low *ξ*. In contrast, since the population-wide strategy is not dependent on infection numbers in any specific region, the population structure is less important and the disease is only rarely completely eliminated from some sub-populations (Supp. Fig. 3). Therefore, a striking reduction in the required restrictions originates from small number fluctuations, the ability of the local strategy to impose restrictions exactly where needed, and its emergent effect of rendering more and more regions disease-free. We find that the weak dependence of restriction time on the leakiness *ξ* for the population-wide strategy and a contrasting threshold-like dependence of the restriction time for the local strategy (Fig. 1d, cf. Supp. Fig. 2) are universal features (across the entire parameter space) that emphasize the benefit of local strategies and the role of cooperativity.

**Figure 2.**
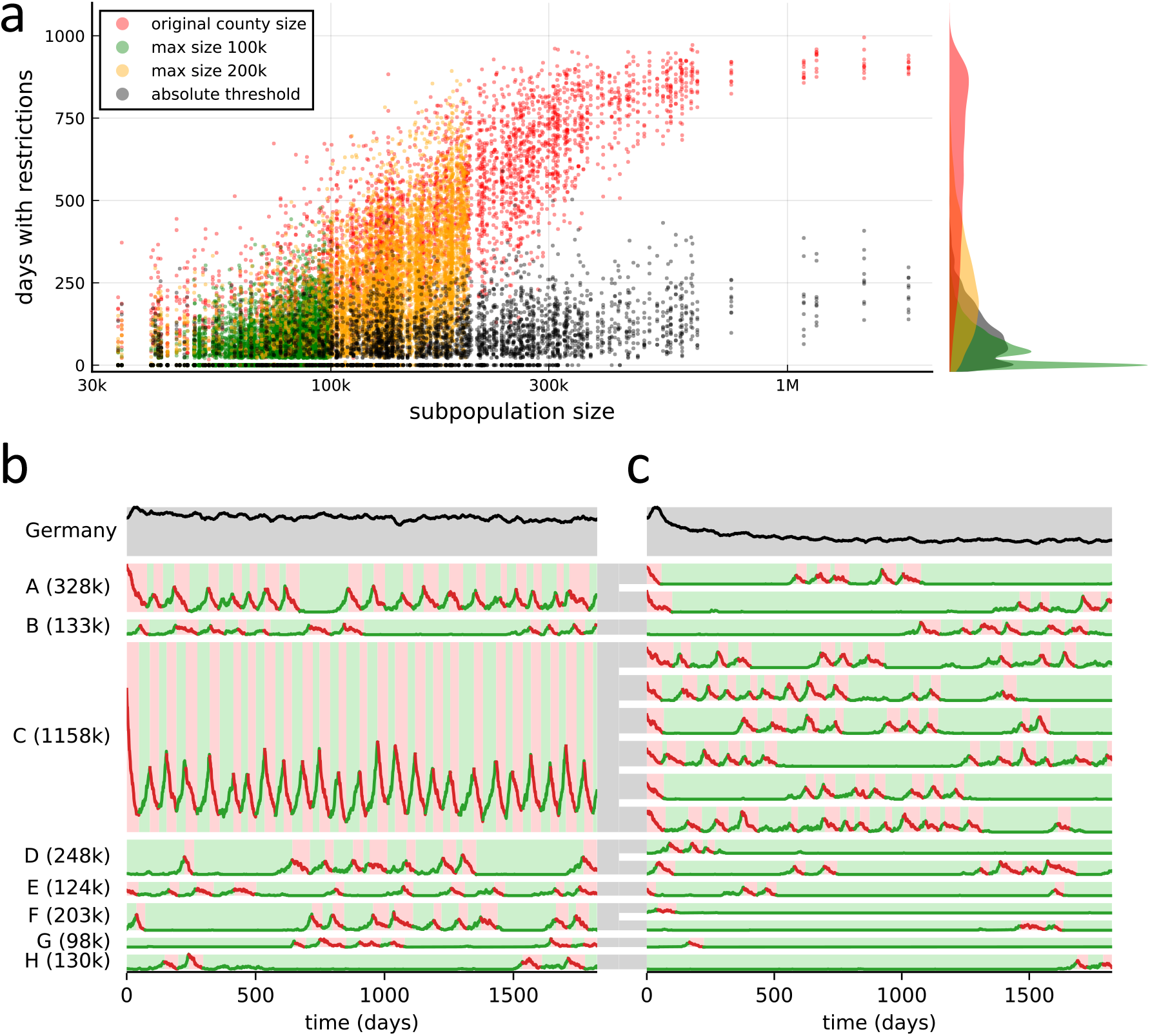
Relative lockdown thresholds cause persistence of the disease in densely populated areas. **a**, Number of days individual sub-populations have to activate severe restrictions because of local outbreaks, plotted against their corresponding size, in simulations of the epidemic over the next 5 years. Scatter plot shows data from 10 realizations of the stochastic dynamics each, for the original German county sizes (red) and further subdivided counties until sub-population sizes of a maximum of 200,000 (yellow) and 100,000 (green) are achieved, all for the same relative lockdown threshold of *θ* = 10 : 100,000. Data for an absolute lockdown threshold of 10 infected individuals for every sub-population regardless of its size is shown in black. The marginal distributions of the time spent under local lockdowns for individuals across the entire population are displayed on the right hand side using the same colors. Remaining parameters: *R*_0_ = 1.29, *ξ* = 1%, *τ*_safety_ = 21. **b**, Dynamics of infection numbers for the case of original county sizes (red data points in panel A) in the first 5 years. Infection numbers in the total German population are shown in the top row. The lower rows show the dynamics in a number of sample counties A-H, with the population size shown in parentheses. Red shading indicates periods with lockdown, green shading indicates no lockdown. **c**, Dynamics in a simulation with large counties further split into equally sized xpulations, such that the maximum sub-population size is 200,000 (yellow data points in panel A). Otherwise, parameters are identical to those in panel B. Gray areas connecting panels B and 2c indicate the split-up.

**Figure 3.**
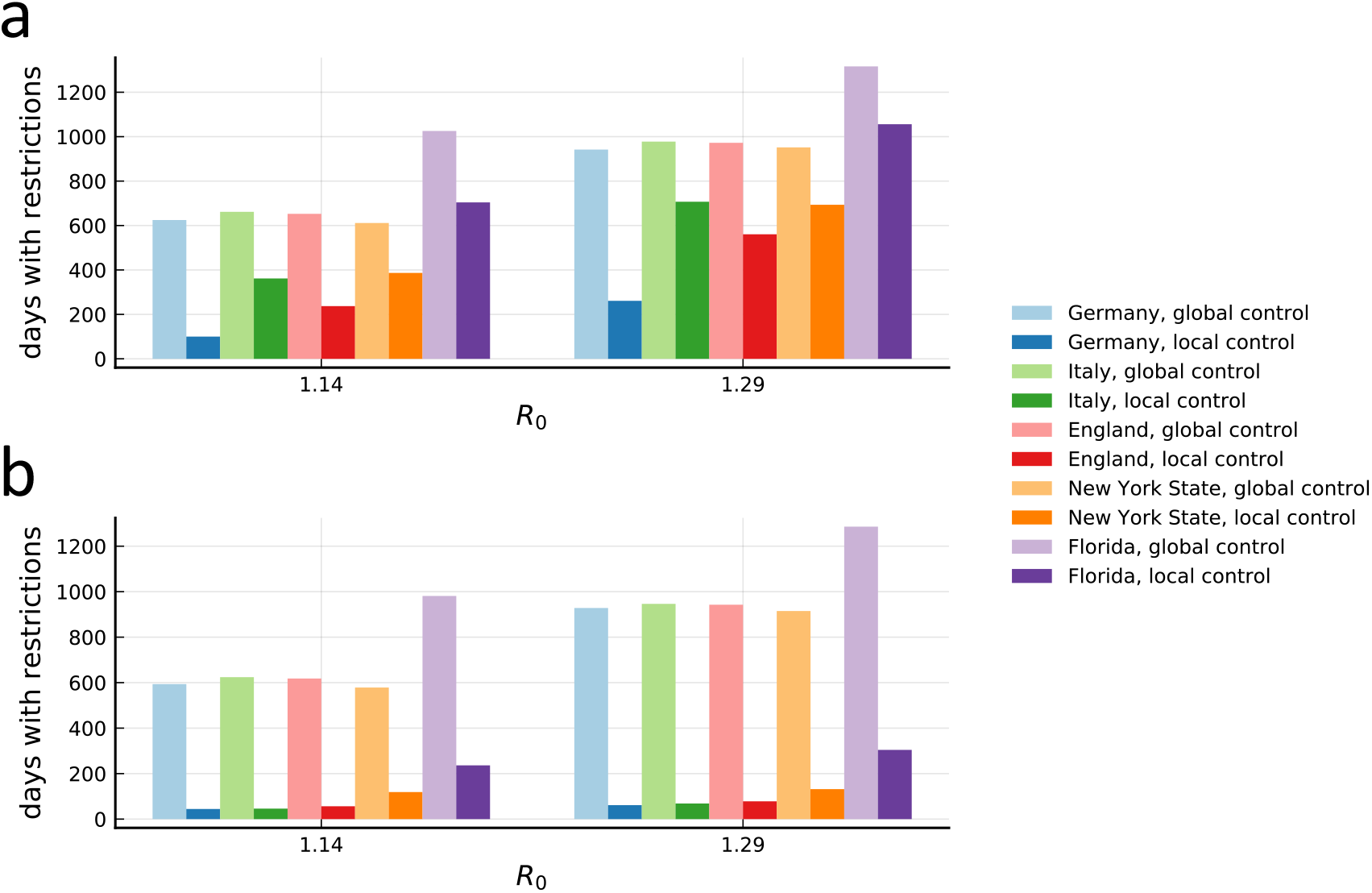
Different county structures lead to varying effectiveness of local measures between countries. **a**, Days with restrictions for the average person for tight global or local control for all five countries included in the analysis with their respective *R*_*l*_ during lockdown (see Supp. Fig. 1). Parameters represent a control strategy with *θ* = 5 : 100,000, *τ*_safety_ = 28 d, a leakiness of *ξ* = 1% and no further subdivision of the counties. For countries with large county sizes, local measures are less effective in bringing down the duration of restrictions required. **b**, Same as in panel A, but for a further subdivision of the counties into sub-populations with a maximum population of 200,000 individuals.

### The role of large sub-population structure

One major difference between individual sub-populations besides their current number of infections is their size. We find that, when the lockdown threshold is defined as a relative proportion *θ* as introduced above, frequent lockdowns are more likely to be required in larger sub-populations. For certain values of the leakiness, *R*_0_ and the lockdown threshold *θ*, these particularly active regions can prevent the overall number of infections from declining, leading to a loss of any advantage that local containment measures can have over a population-wide strategy. For example, using the original sizes of all 412 counties in Germany, there is a clear correlation between the county size and the time span for which the corresponding county has to activate severe restrictions to fight local outbreaks (Fig. 2a, red data).

Densely populated areas prevent the reduction of restrictions in at least two ways: First, the dynamics are characterized by almost periodic switching between lockdown and restriction-free periods (Fig. 2b). The mechanism is similar to the one which leads to the unfavorable performance of the nation-wide lockdown strategy (cf. Fig. 1c), in that absolute case numbers in a sub-population are generally too high to achieve complete elimination of the disease, and so lifting the lockdown even after a long safety margin *τ*_safety_ leads to the immediate resurgence of the epidemic. Besides leading to persistence of the disease in the group itself, this first effect also turns large sub-populations into a continuous source of infections for other regions in the country. Secondly, lockdowns in large-population regions affect a large number of individuals at the same time and thus have a strong impact on the population-wide average time span a person has to live with severe restrictions.

In contrast, if large regions are split up to limit the maximum population size to, e.g., 200,000 people (yellow data in Fig. 2a and Fig. 2c), all sub-populations are given the chance to become intermittently disease-free, avoiding lockdowns for longer periods of time and, cooperatively, leading to a long-term decline of infection numbers in the whole population (compare top rows between Figs. 2b and C). For a maximum size of 100,000 individuals, the effect becomes even stronger (Fig. 2a, green data). For relative thresholds, the further subdivision of large counties effectively leads to smaller absolute thresholds, so it is not surprising that a similar improvement can be observed when regions are not split up, but instead, the same absolute lockdown threshold is used in each sub-population, regardless of its size. This strategy removes the size correlation almost entirely (Fig. 2a, black data) and reduces the average restriction time required to control the epidemic in a similar way. However, it remains to be seen whether such absolute thresholds would be realistic in terms of a practical implementation.

### Differences between countries

Without further subdivision of counties, the effects described above therefore lead to pronounced differences in the reduction that can be achieved in different countries. Countries with larger counties are at a disadvantage when exploiting this natural administrative level for the implementation of a local containment strategy. Using parameters representing a tight control of the epidemic, we therefore find that Germany, with its comparably small counties (7% of the population live in counties with a population larger than 800,000), can substantially reduce restriction time (Fig. 3a) with a local strategy based on a subdivision by county. In contrast, England (25%), Italy (50%), New York State (65%) and Florida (51%) perform considerably worse, with Italy, New York State and Florida not even achieving a 50% reduction. For comparison, when regions are further subdivided into sub-populations of a maximum of 200,000 people, reduction in all countries becomes comparable and substantial (Fig. 3b).

## Discussion

After the successful control of the acute phase, local containment strategies offer a sustainable route for the long-term management of the COVID-19 epidemic. While the exact value of *R*_0_ after all contact and travel restrictions have been lifted is not yet known, our analysis shows that local containment with the same relative lockdown threshold *θ* on average never performs worse than its population-wide equivalent^1^, although parameters that do not achieve an average improvement can lead to an unequal distribution of restrictions across the population (Fig. 1b). However, local containment can lead to a large reduction in the required restrictions for *R*_0_ sufficiently close to one and sufficiently small leakiness. Note that these benefits are based explicitly on discrete, low numbers of infected individuals, similar to effects such as extinction [12] and persistence [10, 11] observed for the ‘free’ evolution of the epidemic. They would therefore not be present in a deterministic mean-field description [3] (see Appendix) as commonly used to track the dynamics of the pandemic.

The importance of entering the ‘small-number regime’ in individual sub-populations becomes evident in at least two ways: First, as we have seen, large sub-populations with the same relative thresholds are less likely to transiently eliminate the epidemic. This is precisely because absolute infection numbers are higher in these sub-populations, and so they fail to benefit from the low number and discreteness of infection events in exactly the same way that the population-wide strategy does. This means that countries such as Germany with its rather small-scale county structure naturally lend themselves to implementing such an approach using existing administrative structures. However, coarse county structure can be compensated for if countries can treat regions over a few hundred thousand inhabitants more strictly or monitor infections (and trigger measures) in less connected smaller sub-populations within those regions separately. The second place where the importance of small numbers shows up is the choice of the relative lockdown threshold. Our results indicate that *θ* around 10: 100,000 achieves desirable results, which are further improved by choosing a large safety margin *τ*_safety_ (Supp. Figs. 5, 6, 7, 8, 9). It is worth putting this *θ* into context: Since we use a mean infectious period of 1/*k* ≈ 7 days based on effective quarantining measures [6, 7], our thresholds can be interpreted as new infections (per population) in the past 7 days. For comparison, a threshold incidence of 50 cases per 100,000 inhabitants in the last 7 days is currently used as an emergency indicator in Germany [9]. Our results therefore suggest that a substantially lower threshold should be employed to get access to the cooperative benefits of local containment measures. Given the current low numbers, this does not seem unrealistic.

In this context, it is interesting to note that, when starting from already low infection numbers, the strength of local control itself is not as critical for the benefit of local measures as it first seems: For a direct comparison, we assumed that, in Germany, instead of *R*_*l*_ = 0.77 during a local lockdown, only a mild reduction in contact rate is achieved, such that the reproductive number is *R*_*l*_ = 0.95, only slightly below 1. For parameters that are otherwise identical to those in Fig. 1, naturally, the absolute restriction time increases (Supp. Fig. 2c) compared to local restrictions with the actual value of *R*_*l*_ (Supp. Fig. 2a). However, there is still a substantial benefit of the local over the global strategy, with a dependence on leakiness that is shifted slightly towards smaller leakiness, similar to that for a higher *R*_0_ (Supp. Fig. 2b). This is similar to the situation of Florida with *R*_*l*_ ≈ 0.9, which could still achieve a reduction in certain parameter regimes (cf. Fig. 3, Supp. Fig. 9). This finding is important for countries which are not willing to apply overly strict measures during a local lockdown.

In the practical implementation, the priority should therefore be on detecting and promptly responding to local increases in case numbers, rather than on an exceedingly strong response. An important contribution is effective testing, essentially reducing the effective lockdown threshold *θ*, which can avoid unnecessary restrictions through a trickle-down effect (see bottom rows in all panels of Supp. Figs. 5, 6, 7, 8, 9), even though in the short run it paradoxically causes more local restrictions. Conversely, less stringent testing leads to a net *increase* in the absolute time restrictions have to be applied and also diminishes the relative improvement of local over global measures. Therefore, insufficient testing might create conditions that make it difficult for the avoidance of unnecessary restrictions as compared to a population-wide strategy to actually materialize, in addition to the general problems it causes [16]. Secondly, in the same way that masks are primarily worn to protect others [17, 18], local containment primarily prevents the spread of the disease to other sub-populations, reducing the average required restrictions for *everyone else*. Cooperation therefore plays a critical role not only on the physical, but also on the political level. Thus, it seems advisable to have national policies in place which define minimum standards for thresholds and local measures to be taken, such that the reaction to local outbreaks is swift and automatic, and not delayed due to political and administrative processes.

An important unknown parameter in our study is the proportion of cross-regional contacts. While related to travel and mobility [19, 20, 21], concrete values for *ξ* are difficult to ascertain. Spatially resolved data on infection chains (for example recorded by the the local health agencies performing the contact tracing) would be of great value in order to quantitatively monitor *ξ* and, if an excessive leakiness is found to impede the efficient management of the epidemic, to implement measures to reduce it. This would only affect cross-region contacts while maintaining freedom for citizens and small businesses within local regions. Note also that a reduction of cross-regional contacts does not necessarily require restrictions in mobility. Rather, policies could aim at reducing their potential infectiousness, which could be achieved, e.g., through frequent testing of the involved personnel or special protective equipment similar to that used in the healthcare setting. As a general principle, our study highlights that the transfer of responsibility to local regions should always go hand in hand with a close monitoring of cross-regional infections in order not to miss out on the benefits.

While first treatment options for COVID-19 are starting to emerge [22] and trials for various vaccine candidates are underway, it is likely that the world will have to live with a simmering epidemic for a while. In our view, thinking about sustainable long-term strategies is indispensable to get through this period with minimal harm. We hope that our study can contribute to this discussion on the basis of rigorous predictions.

## Data Availability

All data and materials to reproduce the findings are either publicly available or contained in the manuscript. While a complete mathematical description of the model is given in the Appendix, the simulation code is available for download at https://gitlab.gwdg.de/LMP-ub/localcontainment.

## Acknowledgments

We are grateful to Viktoryia Novak for graphical design support and thank Michael Wilczek, Viola Priesemann, Jürgen Jost and the remaining Max Planck SARS-CoV-2 task force for stimulating discussions. This work was supported by the Max Planck Society.

## Data availability

All data and materials to reproduce the findings are either publicly available or contained in the manuscript. While a complete mathematical description of the model is given in the Appendix, the simulation code is available for download at https://gitlab.gwdg.de/LMP-pub/localcontainment.

## Appendix

### Mathematical model

#### SIR model with subdivision

We model the epidemic spreading dynamics by considering *n* separate groups or sub-populations with internal SIR dynamics [23] and coupling between the groups. Let *S*_*j*_ and *I*_*j*_ with *j* ∈ 1 … *n* denote the number of susceptible and infected individuals in group *j*, respectively, and *N*_*j*_ the group size. Let

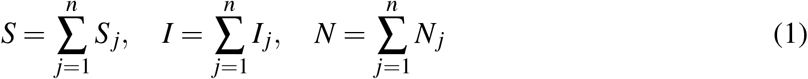

be the corresponding population totals. As in the original SIR model, we assume that individuals have a certain total infectious contact rate *b* = *κν*, where *κ* is the number of contacts with a random other individual per unit time and *ν* is the probability of infection for a contact between a susceptible and an infected individual. We denote by *α*_*j*_(*I*_*j*_, *I*) the probability per unit time for a single susceptible individual in group *j* to become infected, and call this quantity the *infection rate in group j* in the following. The removal of a single infected individual happens with a constant rate, *k*, and can be due to recovery, death or quarantine. We define a *leakiness ξ* to characterize the coupling between local groups: a fraction *ξ* of an individual’s contacts take place with the entire population, whereas the remaining fraction of 1 − *ξ* contacts are restricted to the local group. For this scenario (without containment measures), we therefore have an infection rate for group *j* of

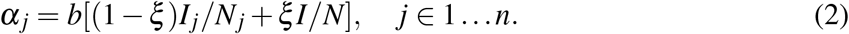

For *ξ* = 0, sub-populations are completely decoupled (no cross-infections between groups occur), whereas for *ξ* = 1, the original SIR model for the population totals *S* and *I* is recovered.

#### Mean field evolution

In a large population with a large number of infected individuals in all sub-populations, deterministic mean field equations capture the dynamics of the epidemic, such that, for all *j* ∈ 1 … *n*

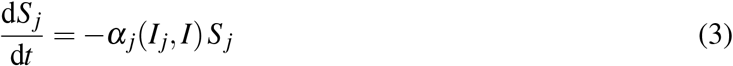

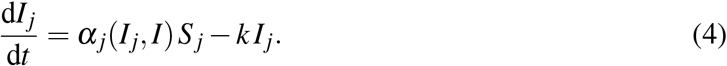

Given a homogeneous initial condition, i.e. with *S*_*j*_/*N*_*j*_ = *S*/*N* and infected *I*_*j*_/*N*_*j*_ = *I*/*N* for all *j, α*_*j*_ reduces to *b I*/*N*, such that the population totals *S* and *I* follow classical SIR dynamics and homogeneity among the groups is preserved, independent of the value of *ξ*. For *ξ* = 1, *α*_*j*_ also reduces to *b I*/*N*, i.e. the subdivision structure becomes irrelevant. Since, by definition, the total infectious contact rate of an individual is always *b*, it is intuitive that the reproductive number is *R*_0_ = *b*/*k* as in the original SIR model, and disease invasion takes place above a critical value of *R*_0_ > 1.

This can be seen rigorously by calculating the average number of infections a single infected individual causes in a fully susceptible population. Let *l* be the group in which the infected individual resides. Then *I*_*l*_ = 1 and *I*_*j*_ = 0 for *j* ≠ *l*. During some time *T*, the number of secondary infections caused directly by this one infected individual therefore is

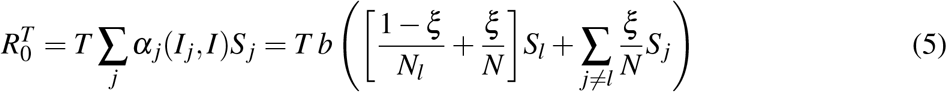

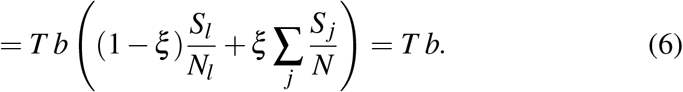

In the last step, we took the large-population limit, where full susceptibility implies *S*_*j*_/*N*_*j*_ → 1 for all *j* (for *j* ≠ *l* the equality holds even for finite populations). With the average duration of a single infection, ⟨*T*⟩ = 1/*k*, we get

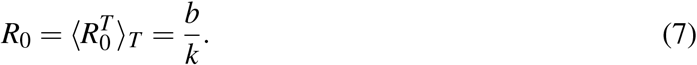

Applying other ‘threshold theorem’ formalisms for subdivided populations [24, 25] leads to the same result.

#### Local lockdown

During a local lockdown, we assume a reduced contact rate of *b*_*l*_ = *q*^2^*b*, where 0 < *q* < 1 is a ‘presence factor’ that indicates the effective availability of an individual for interactions with other individuals during a lockdown. The purpose of this definition of *q* is to be able to associate the reduction in contacts with each class, i.e. susceptibles and infected, individually, as one of them might be in lockdown, while the other one is not, in the case of cross-group infections. During a local lockdown of group *j*, the effective number of susceptible and infected individuals available for contacts will therefore be reduced to *qS*_*j*_ and *qI*_*j*_, respectively. Group-local interactions between them lead to a contribution

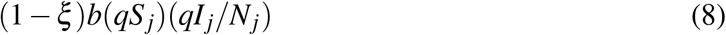

to the infection rate, which simply reduces to (1 −*ξ*)*b*_*l*_*S*_*j*_*I*_*j*_/*N*_*j*_ as expected. To be able to express the effect of local lockdowns on cross-group infections represented by the second term of Eq. (2), we further differentiate between the total number of infected *I*^*l*^ in all groups that are currently in *lockdown* and the corresponding total *I* ^*f*^ in all groups that are currently *free*, with *I* = *I* ^*f*^ + *I*^*l*^. This leads to an overall rate of infections in group *j* of

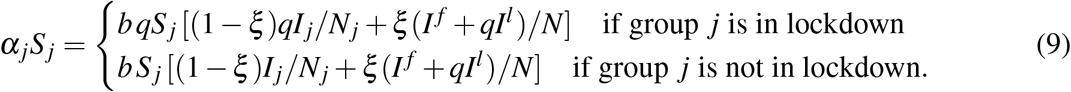

When a group goes into local lockdown, the reduced availability of its susceptible and infected members therefore has two effects: (1) for internal contacts, *b* is reduced to *b*_*l*_, and (2) for cross-infections to/from other groups, the effective rate is reduced and takes an intermediate value between *b*_*l*_ and *b*, which depends on the lockdown state of other groups.

Consequently, if the whole population is in lockdown (*I*^*l*^ = *I, I* ^*f*^ = 0), we have *α*_*j*_ = *b*_*l*_[(1 − *ξ*)*I*_*j*_/*N*_*j*_ + *ξ I*/*N*], identical to Eq. (2), with *b* replaced by *b*_*l*_ as intended. Thus, we can associate the reduced contact rate with a reproduction number

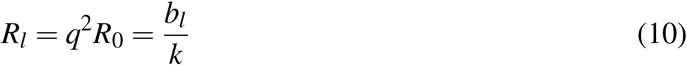

during a population-wide lockdown.

#### Numerical simulation

We carry out exact, stochastic Gillespie simulations [26] of the infection dynamics in the subdivided population, where susceptible and infected individuals in each group are treated as a separate species, i.e. for a total of 2*n* species. The rate for the infection reaction with (Δ*S*_*j*_, Δ*I*_*j*_) = (−1, 1) is *α*_*j*_*S*_*j*_ according to Eq. (9), the rate for the removal reaction with Δ*I*_*j*_ = −1 is *kI*_*j*_. Local lockdown is initiated in group *j* when *I*_*j*_ continuously exceeds the lockdown threshold Θ_*j*_ for 14 days, to account for detection uncertainties and reporting delay (see main text). For relative lockdown thresholds *θ* specified as the infected fraction of the population, we set Θ_*j*_ = *θ N*_*j*_. For absolute thresholds, we set Θ_*j*_ = Θ for all *j* with a universal absolute number of active infections Θ. The lockdown in group *j* is lifted when *I*_*j*_ stays continuously below Θ_*j*_ for a specified amount of time *τ*_safety_ which we call the *safety margin*. While *τ*_safety_ has a certain minimum value because of the same unavoidable processes that delay the detection of a local outbreak and initiation of lockdown, further increasing *τ*_safety_ can be an element of the containment strategy and is therefore up to the policy makers. For the population-wide lockdown strategy, the threshold and delay criteria are applied to the total number of infected individuals *I* and lockdown is initiated/lifted in all groups simultaneously.

#### Data sources and parameter estimation

##### Estimation of *R*_*l*_

During the initial period of the pandemic, many countries enforced a nation-wide lockdown for multiple weeks. Infection data for this time frame allows for a straight forward estimation of the reproduction factor *R*_*l*_ with mitigation strategies in place. We employ the software *EpiEstim* [27] along with daily case data (up until June 12, 2020) for a time-resolved estimate of the reproduction number. For Germany, the infection data is retrieved from the Robert-Koch-Institute [28] using the REST api [29]. Data for Italy and England were available on respective government websites [30, 31] and for the US we used data provided by the Johns Hopkins University [1]. We impose a distribution over the serial interval for COVID-19 [32] to make use of the *EpiEstim* method *uncertain_si*. For daily estimates of *R* a 14-day interval is considered. As is illustrated in Supp. Fig. 1, we average over the ten lowest daily estimates to extract a reasonable value for *R*_*l*_.

##### Initial conditions and group sizes

To simulate approximations of real countries we gather official census data on a county level resolution for England [33], Italy [34], the US [35] and Germany [36] to set up the sub-populations. These data are then combined with the infection numbers (as of June 12, 2020) to compute the current sizes of the susceptible and infected parts of the population in each county. Where no information about recoveries is available we approximate the number of active cases to be equal to all new infections reported within the previous 14 days (a conservative estimate given our assumed infectious time of 1/*k* = 7 days, see main text). For more fine-grained control when investigating the size-dependent effects in the mitigation strategy, the counties are optionally further subdivided such that no individual group exceeds a certain number of individuals. Infected individuals are randomly assigned to one of the equally sized sub-groups in the splitting process.

---

**Supplementary Figure 1.**
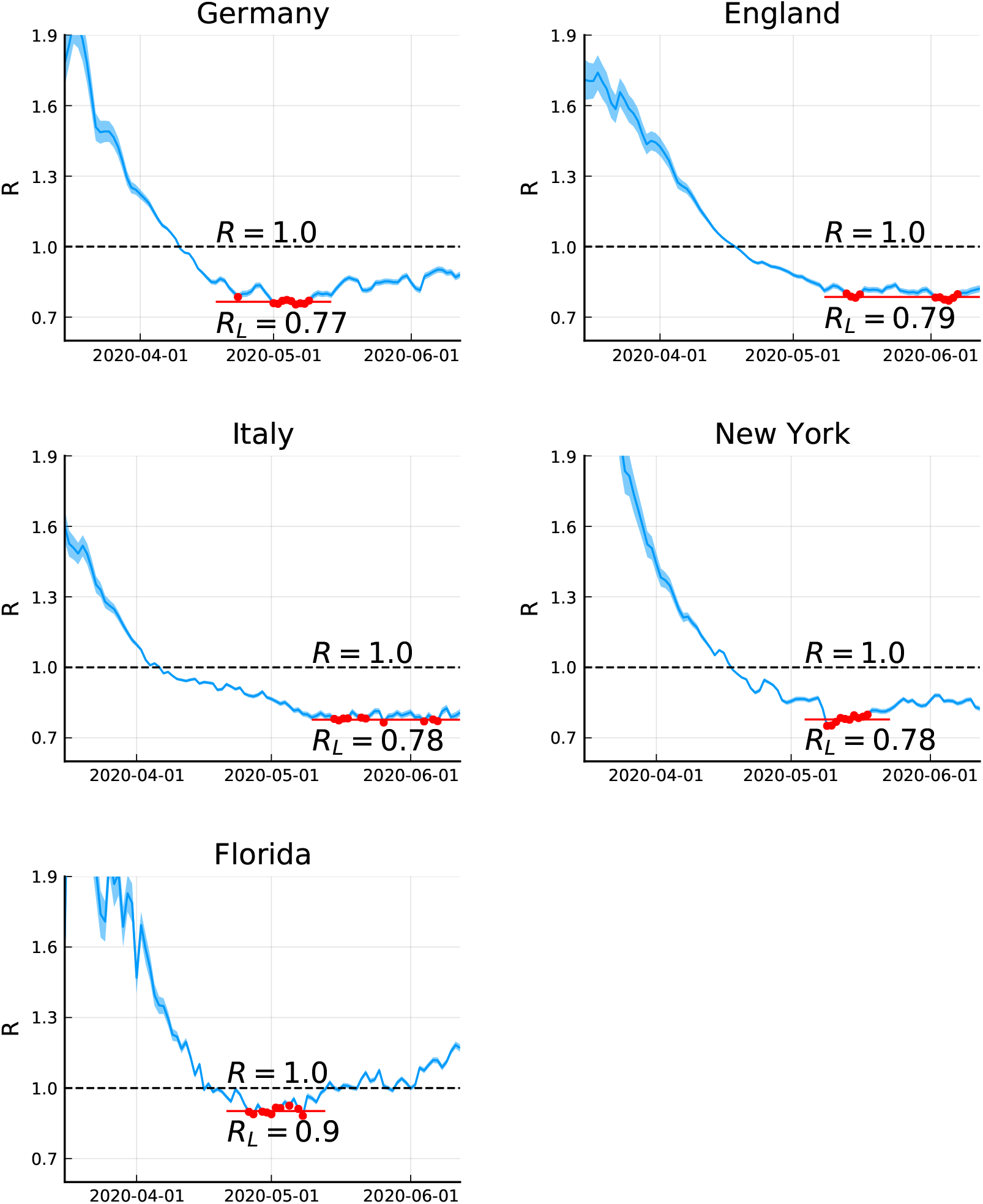
Estimation of the reproduction number during lockdown. Time evolution of 14-day estimates [27] for the reproduction number of COVID-19 in different locations. In all cases, the reproduction number drops as mitigation strategies are put in place. The ten lowest daily estimates averaged to estimate *R*_*l*_, the effective reproduction factor during lockdown, are highlighted in red.

**Supplementary Figure 2.**
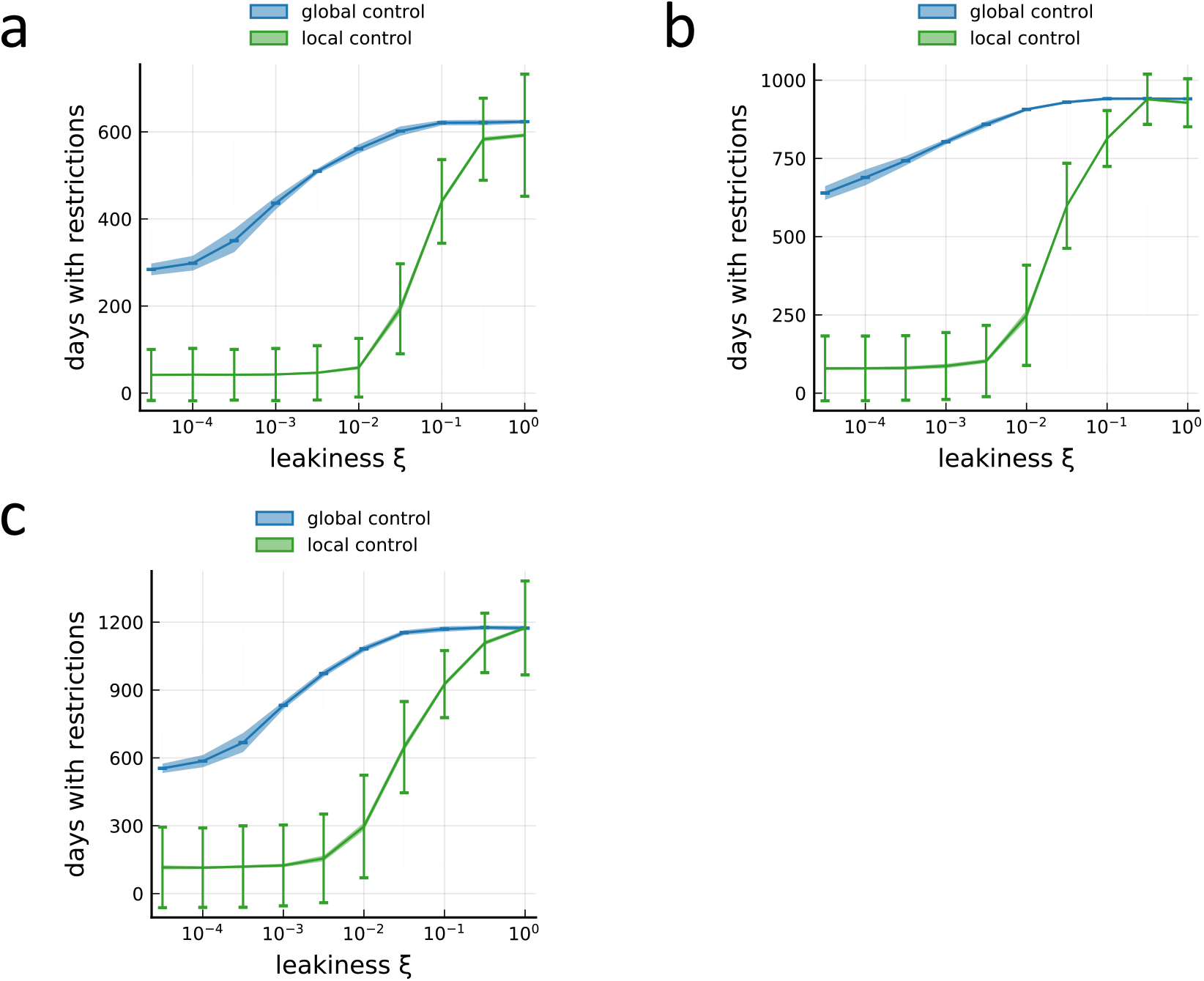
Reducing leakiness causes a transition to low required restriction time for local measures. **a**, Number of days the average person in the population will have to spend in lockdowns within the next 5 years for the full spectrum of leakiness values, using the county structure and current active case numbers from Germany. Reproduction of Fig. 1d. Parameters identical to those used in Fig. 1b for the lower value of *R*_0_ = 1.14. Error bars indicate standard deviation across members of the population in a single simulation (averaged across 20 realizations), shading around lines indicates standard deviation of the average across realizations. **b**, The same as in panel A, but for the higher value of *R*_0_ = 1.29 in Fig. 1b. **c**, The same as in panel A, but for *R*_*l*_ = 0.95 during lockdown, substantially higher than the value achieved during the actual country-wide lockdown in Germany (cf. Supp. Fig. 1).

**Supplementary Figure 3.**
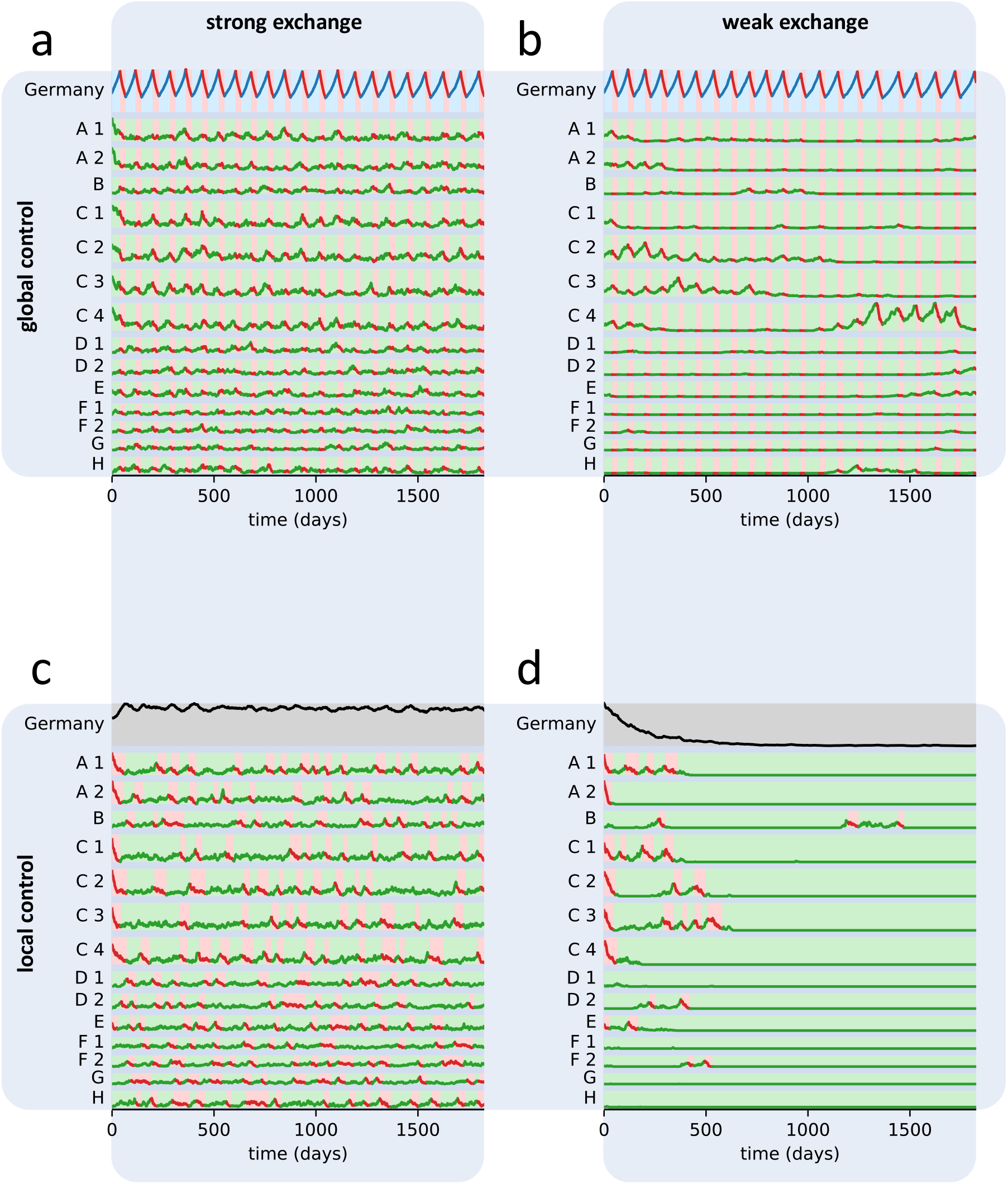
Evolution of infections individual sub-populations for different strategies. Parameters identical to those shown in Fig. 1b, for the lower value of *R*_0_. **a**, Population-wide control and strong exchange (*ξ* = 32%). **b**, Population-wide control and weak exchange (*ξ* = 1%). **c**, Local control and strong exchange (*ξ* = 32%). **d**, Local control and weak exchange (*ξ* = 1%).

**Supplementary Figure 4.**
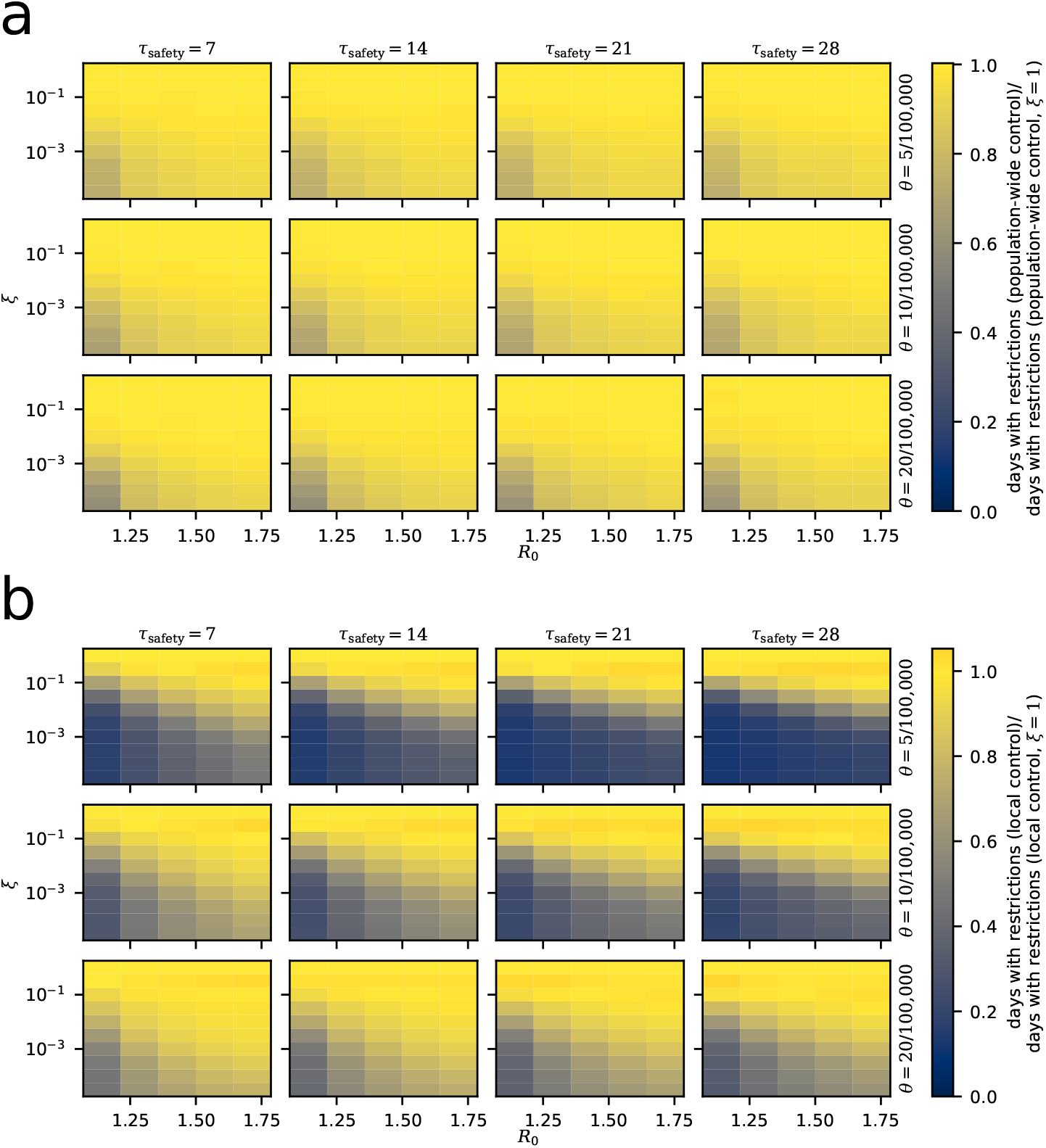
Absolute comparison of different strategies for Germany. All data represent averages from 20 simulations. **a**, Number of days the average person in the population will have to spend under restrictions within the next 5 years under the population-wide control strategy, using the county structure and current active case numbers from Germany without further subdivision. Data normalized by the data for *ξ* = 1, i.e. a fully interconnected population without subdivision. **b**, Same as in panel A, but for the local control strategy.

**Supplementary Figure 5.**
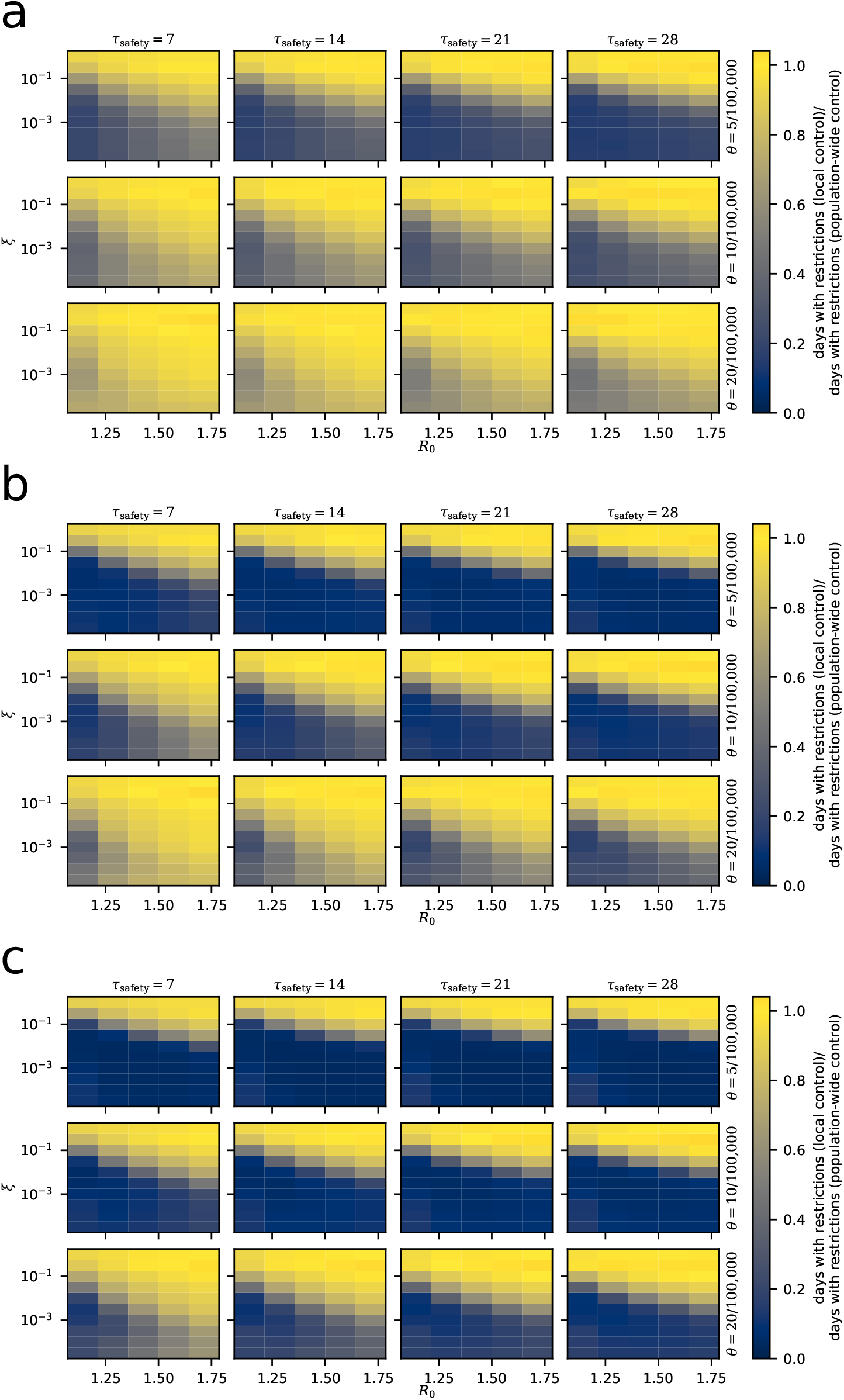
Strategy comparison for Germany for all simulated parameters. All data represent averages from 20 simulations. **a**, Number of days the average person in the population will have to spend under restrictions within the next 5 years under the local control strategy, using the county structure and current active case numbers from Germany without further subdivision. Data normalized by the corresponding values for population-wide control of restrictions. **b**, Same as in panel A, but with counties further subdivided into sub-populations of at most 200,000 **c**, Same as in panel A, but with counties further subdivided into sub-populations of at most 100,000

**Supplementary Figure 6.**
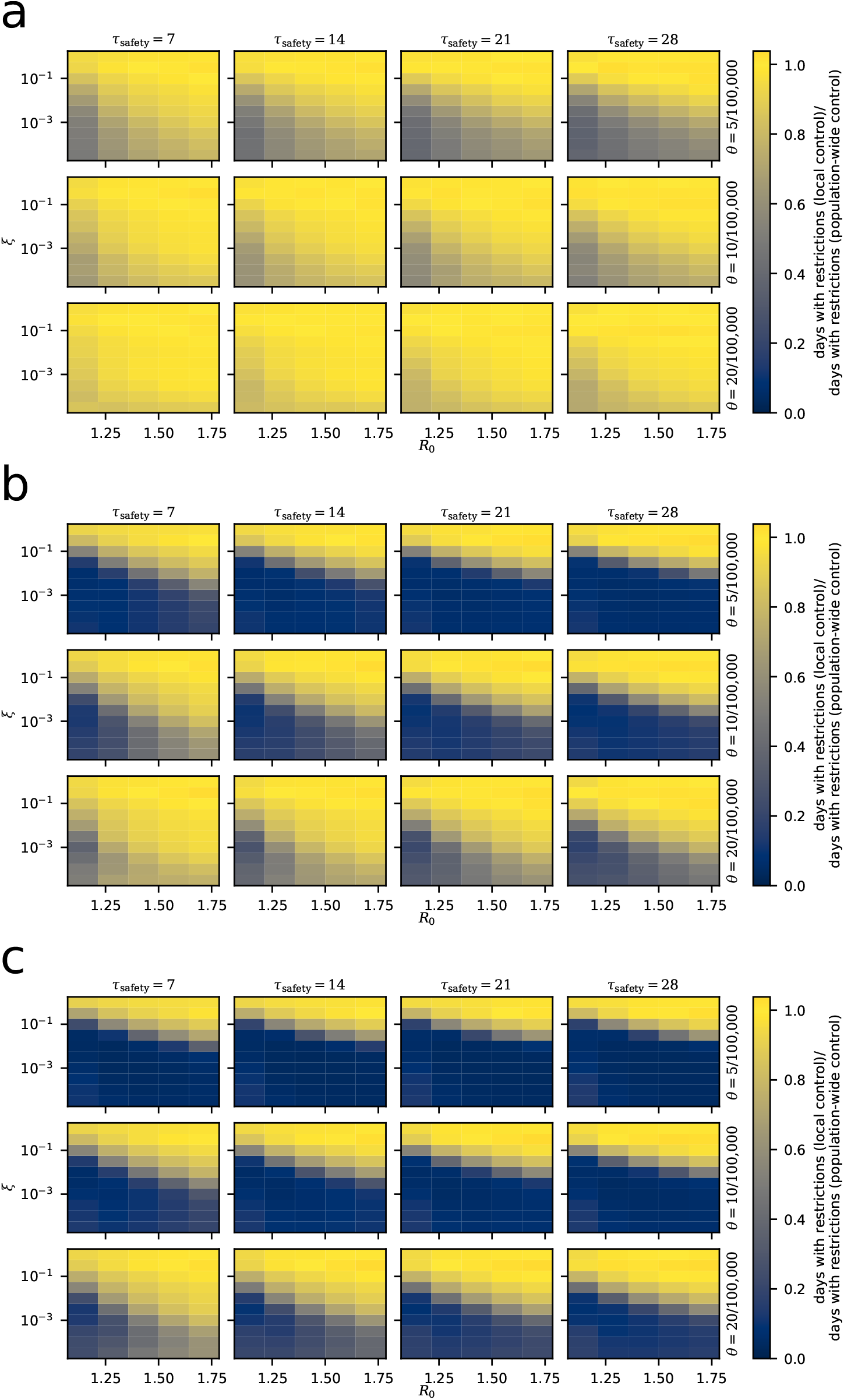
Strategy comparison for Italy for all simulated parameters. Otherwise as in Supp. Fig. 5. **a**, without subdivision **b**, 200k subdivision1**c**, 100k subdivision

**Supplementary Figure 7.**
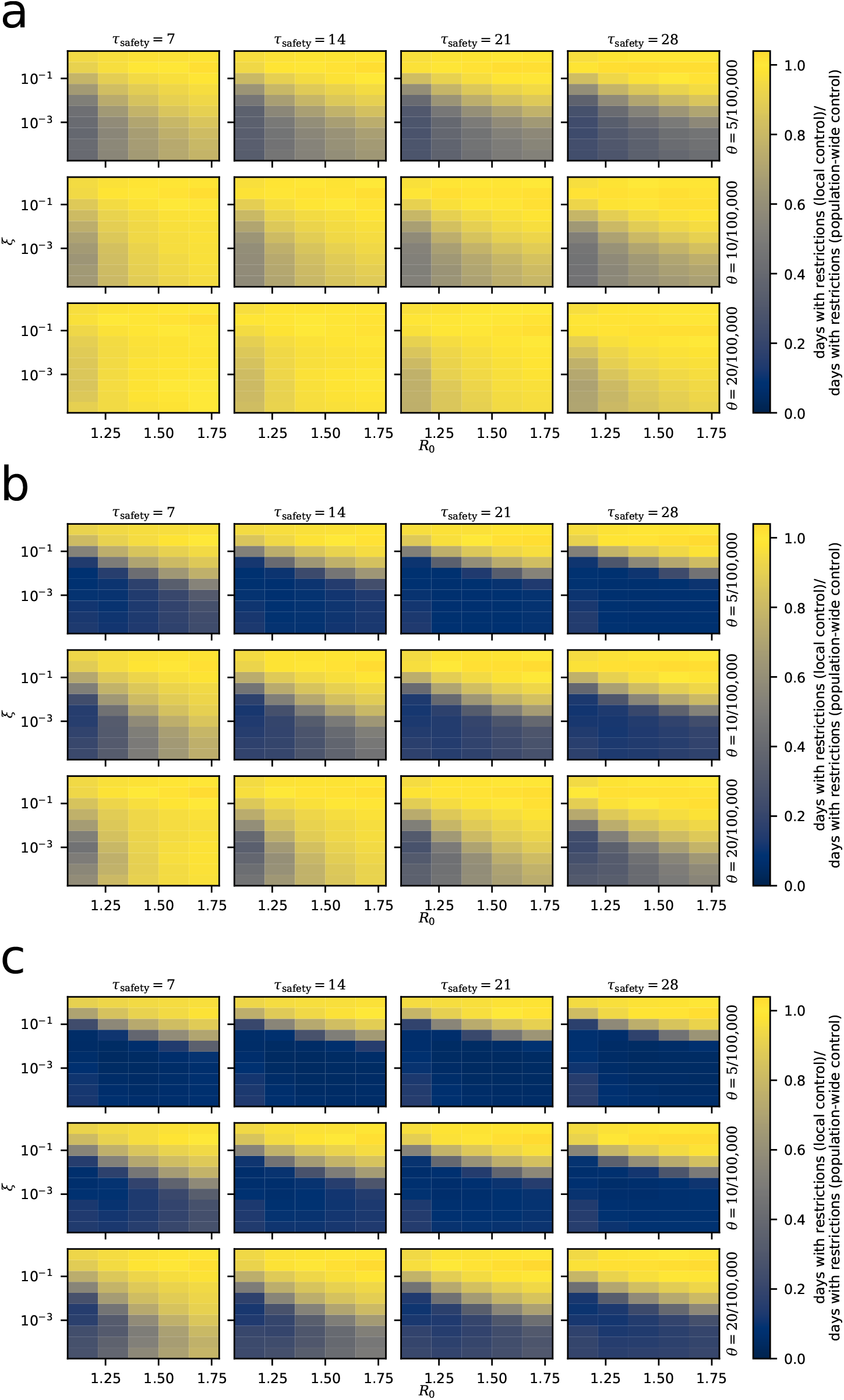
Strategy comparison for England for all simulated parameters. Otherwise as in Supp. Fig. 5. **a**, without subdivision **b**, 200k subdivision **c**, 100k subdivision

**Supplementary Figure 8.**
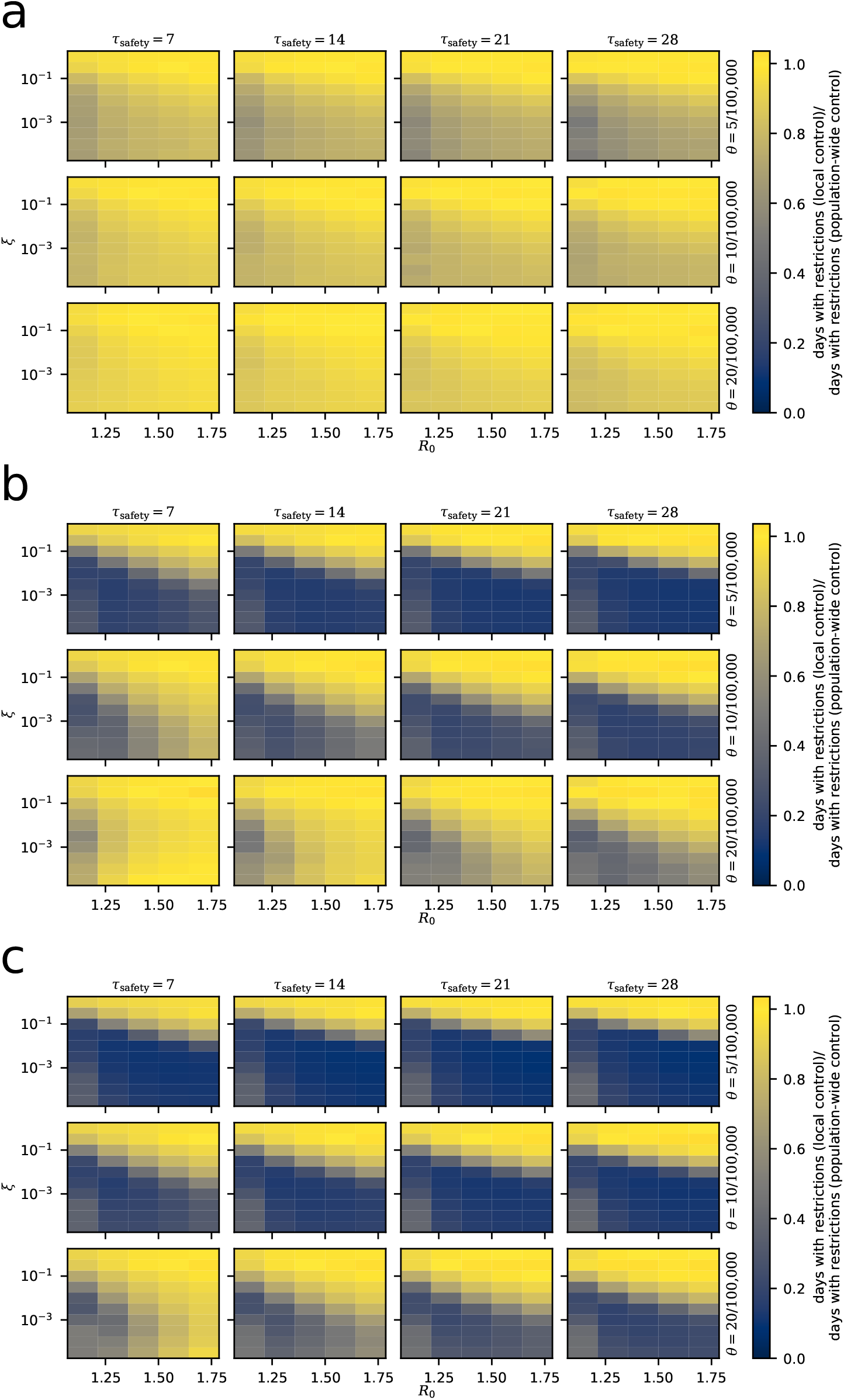
Strategy comparison for New York State for all simulated parameters. Otherwise as in Supp. Fig. 5. **a**, without subdivision **b**, 200k s1ubdivision **c**, 100k subdivision

**Supplementary Figure 9.**
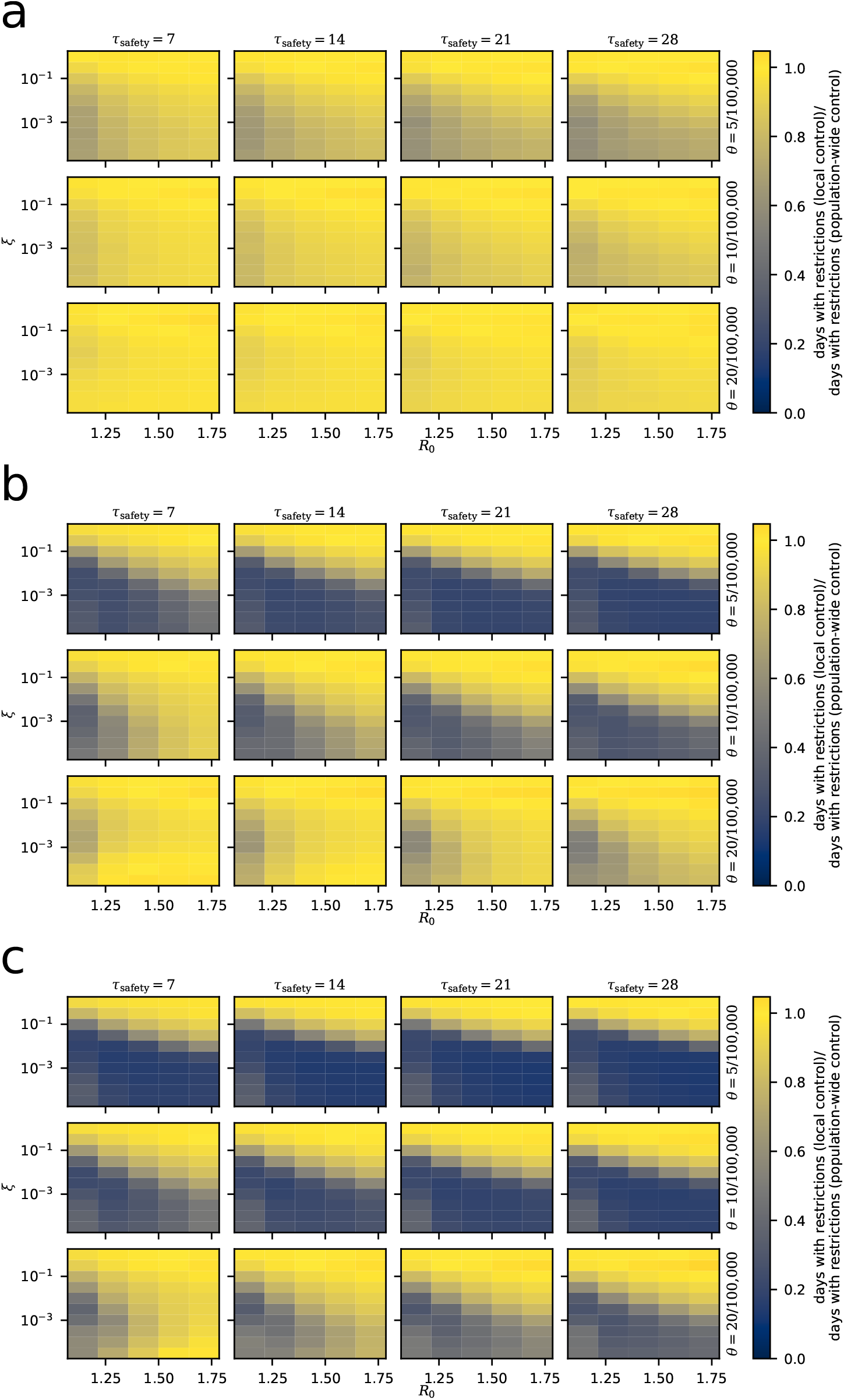
Strategy comparison for Florida for all simulated parameters. Otherwise as in Supp. Fig. 5. **a**, without subdivision **b**, 200k subdivision1**c**, 100k subdivision

**Supplementary Figure 10.**
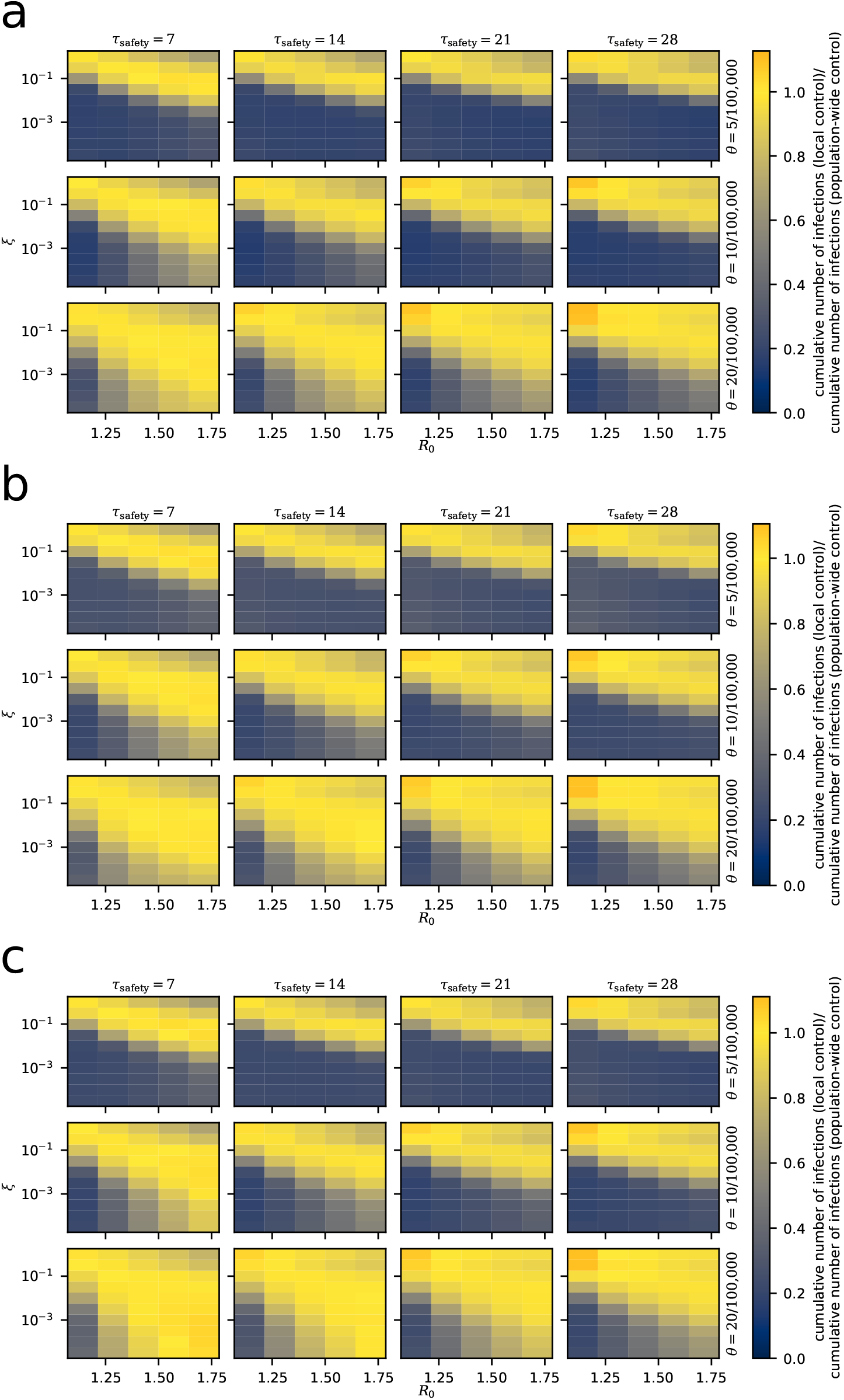
Cumulative infections after 5 years. Values obtained from the local strategy are normalized by the corresponding measurement using the population-wide strategy. All data represent averages from 20 simulations. **a**, Germany **b**, Italy **c**, England

**Supplementary Figure 11.**
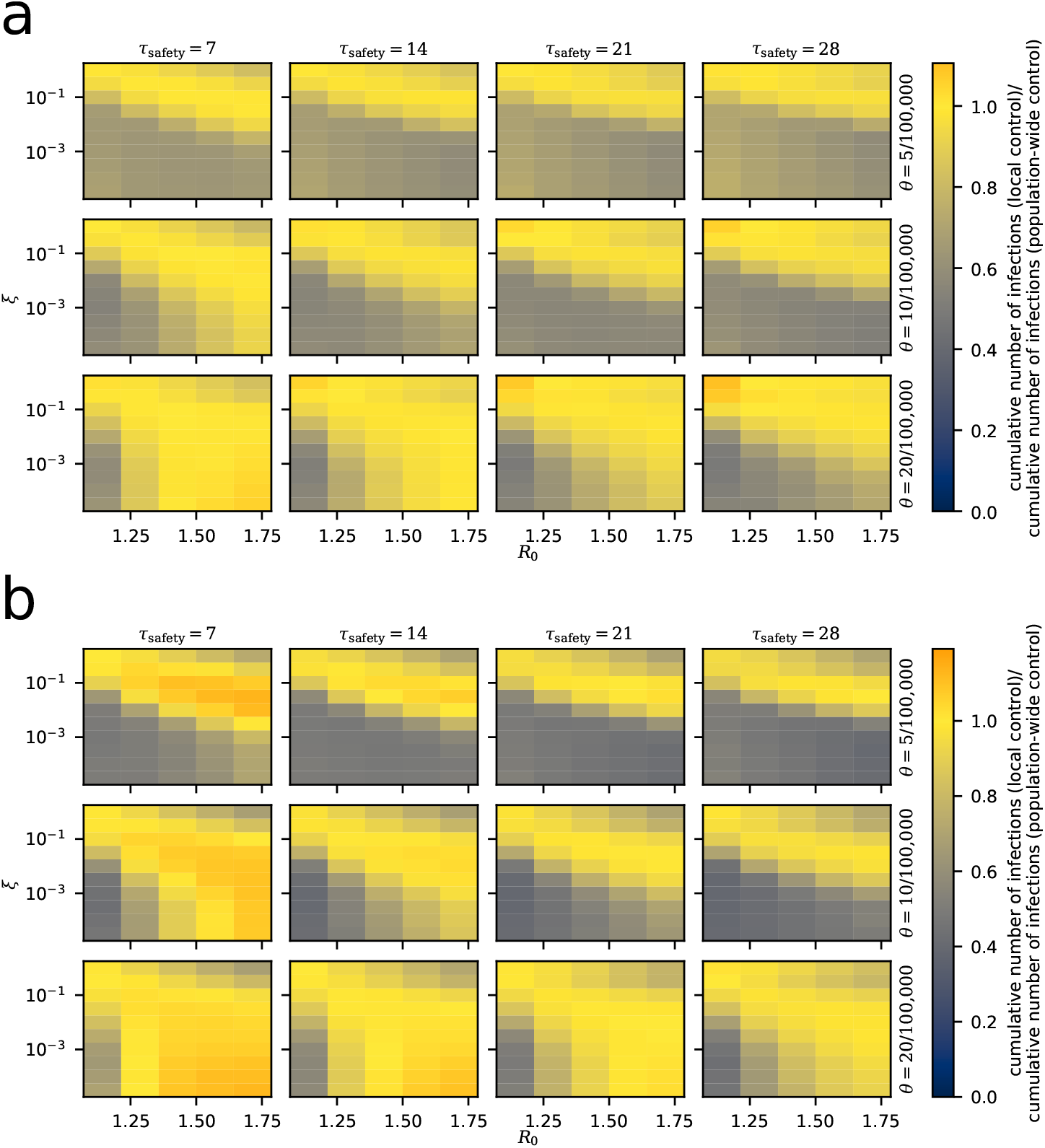
Cumulative infections after 5 years. As in Supp. Fig. 10, but for **a**, New York State **b**, Florida

## Footnotes

1 except for a slight increase in the cumulative number of infections for particular parameters, see Supp. Figs. 10, 11

## Notes

### Competing Interest Statement

The authors have declared no competing interest.

